# Multidrug therapy with terbinafine plus daily fluconazole is more effective than terbinafine alone or terbinafine plus weekly fluconazole in current epidemic of altered dermatophytosis in India: Results of a randomized pragmatic trial

**DOI:** 10.1101/19005025

**Authors:** Sanjay Singh, Bandana Jha, Prakriti Shukla, Vinayak N Anchan

## Abstract

**Background:** Treatment responsiveness of tinea has decreased considerably in recent past in India. We tested effectiveness of oral terbinafine daily plus fluconazole weekly (TFw) and terbinafine daily plus fluconazole daily (TFd) versus oral terbinafine daily (T) in tinea corporis, tinea cruris and tinea faciei in a pragmatic randomized open trial.

**Methods:** One hundred and seventeen microscopy confirmed patients were allocated to T (6 mg/kg/day), TFw (terbinafine 6 mg/kg/day+fluconazole 12 mg/kg once weekly), or TFd (terbinafine 6 mg/kg/day+fluconazole 6 mg/kg/day) groups by concealed randomization and treated for 8 weeks or cure. Each group included 39 patients.

**Results:** At 4 weeks, 9 (23.1%), 8 (20.5%) and 14 (35.9%) patients were cured in T, TFw and TFd groups, respectively (P=0.279). At 8 weeks, number of patients cured was as follows: T 13 (33.3%), TFw 18 (46.2%) and TFd 25 (64.1%). TFd was more effective than T (P=0.012), other comparisons were not significantly different. However, effect size as calculated by number needed to treat (NNT) (versus terbinafine) was 8 for TFw and 4 for TFd. Relapse rates one month after cure were similar in all groups (P=0.664).

**Conclusions:** In view of cure rates and NNT, terbinafine plus daily fluconazole is more effective than terbinafine alone or terbinafine plus weekly fluconazole in current epidemic of altered dermatophytosis in India.

**One Sentence Summary:** Terbinafine plus daily fluconazole is more effective than terbinafine alone or terbinafine plus weekly fluconazole in current epidemic of altered dermatophytosis in India.

## Introduction

Unprecedented changes in the epidemiology, clinical features and treatment responsiveness of tinea infections have been noted in recent past in India.^1-4^ Recent data show that effectiveness of oral terbinafine,^5, 6^ fluconazole, itraconazole and griseofulvin has decreased considerably, even with 8 weeks of treatment.^6^ Decreased effectiveness of terbinafine has been attributed to mutation in the squalene epoxidase gene.^7,8^ Importance of finding effective methods of treating tinea cannot be overemphasized.

In the present study, we tested the hypothesis that using a regimen consisting of more than one oral antifungal drugs may produce better treatment outcome. We performed a three-arm pragmatic randomized open trial comparing effectiveness of oral terbinafine daily (T, active control) versus two experimental regimens, terbinafine daily plus fluconazole weekly (TFw) and terbinafine daily plus fluconazole daily (TFd).

## Methods

### Setting

The study (registered with Clinical Trials Registry-India, registration number CTRI/2017/12/010770) was conducted at a tertiary care centre after obtaining approval from the Ethics Committee of the Institute of Medical Sciences, Banaras Hindu University.

### Sample size

A sample size of 35 patients in each group was determined using expected cure rates of 30% and 70% with the control and experimental regimens with type I error rate of 0.05, type II error rate of 0.1 and dropout rate of 20%.^9^

### Selection criteria and enrolment

Patients with tinea corporis, tinea cruris or tinea faciei or a combination of these conditions were included in the trial. Inclusion criteria were (a) clinical diagnosis of tinea corporis, tinea cruris or tinea faciei, (b) microscopic confirmation (potassium hydroxide [KOH] microscopy), (c) age 4 to 80 years, and (d) no treatment with terbinafine or fluconazole in last one month. Exclusion criteria were (a) presence of any other type(s) of tinea, e.g., onychomycosis, (b) pregnancy, (c) lactation, (d) inability to come for follow-up, (e) history of adverse reaction to terbinafine and/or fluconazole, (f) abnormal baseline investigations (complete blood count [CBC], liver and renal function tests [LFT, RFT]), (g) current treatment with drugs likely to cause interaction with terbinafine and/or fluconazole, and (h) history of renal, liver or cardiac disease or chest pain. A witnessed, written and informed consent was given by the patients, or by a parent in case of minor patients. All female patients of child bearing age were advised to avoid pregnancy during treatment.

Five hundred and fifty-seven patients with clinical diagnosis of tinea corporis, tinea cruris or tinea faciei or a combination of these conditions attending dermatology outpatient department were assessed for eligibility. A total of 117 patients satisfied the selection criteria and were assigned to one of the three treatment groups by block randomization on the basis of random numbers generated online using Research Randomizer (https://www.randomizer.org/). Each treatment group comprised 39 patients (Figure 1) (allocation ratio 1:1:1). Patients were enrolled from September 2017 to November 2017. Allocation was concealed using sequentially numbered, sealed, opaque envelopes, which contained the allocation code written on a folded paper, the envelopes were opened after enrolment of the patients. Both random sequence generation and allocation concealment were done by a person unrelated to the trial.

**Figure 1.**
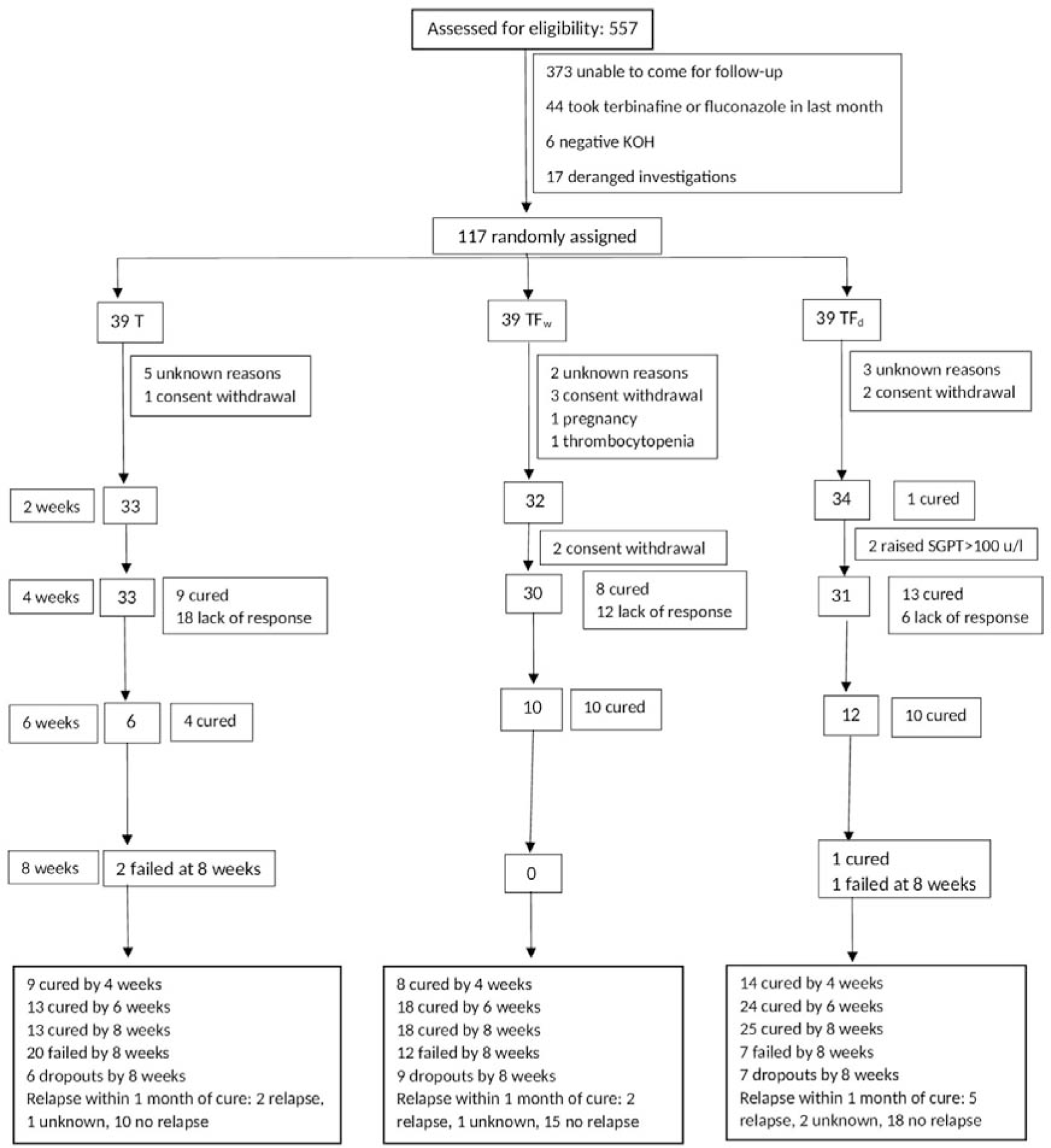
Flow diagram of the progress of the trial through its different phases. SGPT, serum glutamic pyruvic transaminase enzyme.

### Interventions

Dosing schedule of terbinafine was 6 mg/kg/day in all treatment groups. Patients in the TFw group, in addition, received fluconazole 12 mg/kg once a week. To ensure good compliance, a day of the week was fixed for fluconazole intake for this group of patients. Patients in the TFd group received fluconazole 6 mg/kg/day along with daily terbinafine. Patients were advised against using any other treatment.

### Follow-up

Following data were recorded for each patient: body surface area (BSA) affected, treatment given, investigations (CBC, LFT, RFT) at baseline, followed by CBC and LFT every 2 weeks, microscopy results, and follow-up data. The most severe lesion was identified as the index lesion, from which scraping and KOH microscopy was performed at baseline and then at every subsequent visit. Patients were followed up at 2 weekly intervals up to a maximum of 8 weeks or cure, whichever occurred earlier.

### Measurement of treatment effect

Cure was defined as occurrence of both clinical cure (complete clearance of lesions) and microscopic cure (negative KOH microscopy). Presence of post-inflammatory hyperpigmentation at the site of healed tinea was not considered a feature of tinea. Any of the following was considered treatment failure (a) no improvement or worsening of the disease in 4 weeks in patient’s assessment, (b) presence of skin lesions at 8 weeks, and (c) positive microscopy at 8 weeks. Patients who were cured were asked whether they were fully satisfied with the treatment outcome or not. Patients in whom the treatment failed were treated with oral itraconazole outside the clinical trial.

### Measurement of compliance

Patients were asked to bring used strips of tablets and the strips in use at each follow up visit to assess compliance. Compliance was considered to be good if a patient had taken 80% or more of the prescribed number of tablets, and poor if less than 80% tablets were taken during treatment.

### Assessment for relapse

Those patients who were cured were assessed again 1 month after the cure to look for relapse of tinea.

### Outcome measures

Number of patients cured at 8 weeks with the different regimens was the primary outcome measure. Secondary outcome measures included number of patients cured at 4 weeks, relapse rate, and frequency of adverse events. In addition, severity of itching was noted on a 0 to 3 scale (0, none; 1, mild; 2, moderate; 3, severe) at baseline and subsequent follow-ups.

### Statistical analysis

For the baseline variables, mean and standard deviation (SD) or median and interquartile range (IQR) were calculated depending on the distribution of data. Number of patients who had used topical steroid, cure rates, relapse rates and compliance were compared by Fisher exact test. P values less than 0.05 were considered significant. All P vales are two-tailed. Absolute risk reduction (ARR) of treatment failure versus the control (terbinafine daily) and number needed to treat (NNT) were calculated. When appropriate, 95% confidence intervals (CI) were calculated. Intention to treat analysis was performed for the efficacy data. Compliance data were analysed on per protocol basis. Denominator for calculating compliance was the total number of patients who were cured and those who came for follow ups up to 8 weeks but were not cured. Denominator for calculation of relapse rate was the number of patients who had achieved cure. For calculating relapse rate, those cured patients who could not be followed up were considered relapsed.

## Results

### Baseline characterises of the patients

Patients in the 3 groups were comparable at baseline with regard to age, duration of tinea, and body surface area affected (Table 1).

**Table 1.** Baseline characteristics of the patients

|  | T (n=39) | TFw (n=39) | TFd (n=39) |
| --- | --- | --- | --- |
| Age, years<br>(mean±SD) | 33.9±15.2 | 30.1±14.0 | 27.8±13.1 |
| Female (%) | 25.6 | 38.5 | 10.3 |
| Duration, months,<br>median (interquartile<br>range) | 3 (1-6.5) | 3.5 (1-7.5) | 3 (1-6) |
| BSA affected, %, median (interquartile<br>range) | 2 (2-5.5) | 2 (1-3.5) | 3 (2-4) |
| Co-morbidities | Hypertension 1<br>Diabetes mellitus 2<br>Epilepsy 1<br>Arthritis 1<br>Scalp psoriasis 1<br>Urticaria 1 | Urticaria 1<br>Xanthelasma 1<br>Vitiligo 1 | Diabetes mellitus 1<br>Urticaria 1<br>Asthma 1<br>PLE 1<br>Scalp psoriasis 1<br>BPH 1 |

T, terbinafine daily; TFw, terbinafine daily and fluconazole weekly; TFd, terbinafine daily and fluconazole daily; BPH, benign prostatic hypertrophy; BSA, body surface area; PLE, polymorphic light eruption; SD, standard deviation.

Fifty-three patients had tinea corporis et cruris, 24 tinea corporis, 19 tinea corporis et cruris et faciei, 10 tinea cruris, 6 tinea faciei, 4 tinea corporis et faciei, and 1 patient had tinea cruris et faciei. Fifty patients had received some treatment in past 1 month, these included topical steroid combinations by 25 patients (8, 8, and 9 patients in T, TFw, and TFd, respectively, P=0.999), oral itraconazole by 9 patients, topical antifungal and topical salicylic acid by 6 patients each, and oral steroid by 4 patients.

### Cure rates at 4 weeks

At 4 weeks, 9 (23.1%), 8 (20.5%) and 14 (35.9%) patients were cured in T, TFw and TFd groups, respectively (Figure 1) (intention to treat analysis). The cure rates were not significantly different (P=0.279).

### Cure rates at 8 weeks

Thirteen (33.3%, 95% CI 20.6 to 49.1), 18 (46.2%, CI 31.6 to 61.4) and 25 (64.1%, CI 48.4 to 77.3) patients were cured at 8 weeks in T, TFw and TFd groups, respectively (P= 0.026) (Figure 1) (intention to treat analysis). Comparisons of different pairs of treatment groups showed that TFd was more effective than T (P=0.012), while other pairs were not significantly different with regard to cure rates at 8 weeks (Table 2).

**Table 2.**
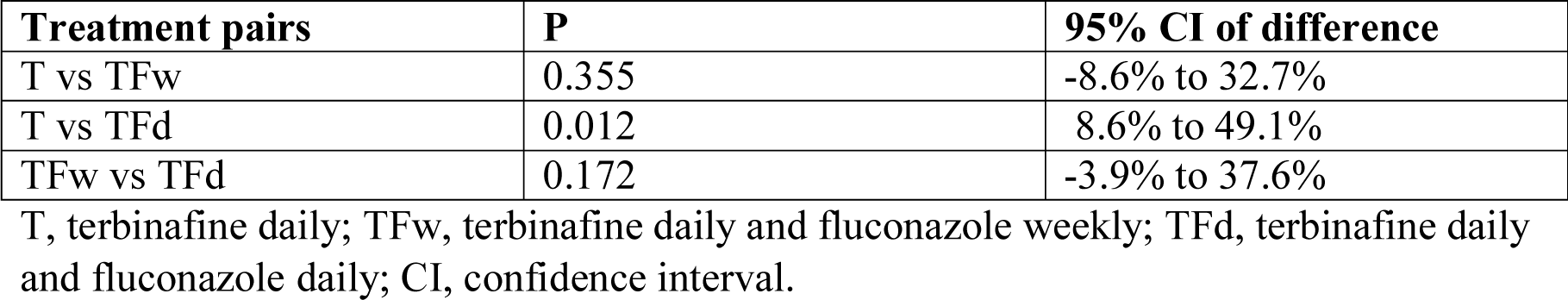
Comparison of the cure rates at 8 weeks with the three treatments

### Severity of itching

Severity of itching decreased in all treatment groups as the treatment progressed and itching was absent in all patients at the time of cure (Table 3).

**Table 3.** Severity of itching

|  |  | Baseline | 4 weeks | 8 weeks |
| --- | --- | --- | --- | --- |
| All patients | T | 2.33±0.762 | 1 (0-3) | 1 (1.25-1.75) |
|  | TFw | 2.38±0.771 | 1 (0-2) | 1 (1-1) |
|  | TFd | 2.35±0.767 | 0 (0-1) | 1 (0.5-1.5) |
| Patients cured at 4 weeks | T | 2.1±0.83 | 0±0 |  |
|  | TFw | 2.25±0.82 | 0±0 |  |
|  | TFd | 2.28±0.79 | 0±0 |  |
| Patients cured at 8 weeks<br>(excluding those cured at 4 weeks) | T | 3±0 |  | 0±0 |
|  | TFw | 2.44±0.83 |  | 0±0 |
|  | TFd | 2.45±0.65 |  | 0±0 |

Normally distributed data are presented as mean±SD and the data which are not normally distributed is presented as median and interquartile range (IQ1-IQ3).

### Relapse rates

Three out of 13 cured patients (23%, 95% CI 7.5 to 50.9), 3 of 18 (16.7%, CI 16.7, 5.0 to 40.1) and 7 of 25 (28%, CI 14.1 to 47.8) patients relapsed within a month of cure in the treatment groups T, TFw and TFd, respectively (Figure 1). Relapse rates were not significantly different among the 3 groups (P=0.664).

### Compliance

Thirty out of 33 patients, 21 of 30, and 29 of 32 patients had good compliance in the T, TFw, and TFd groups, respectively. The difference was not significant (P= 0.053).

### Patients’ satisfaction with treatment outcome

All patients who were cured were fully satisfied with the outcome.

### Number needed to treat

Number needed to treat (NNT) for the experimental group TFw was 8 and for TFd it was 4 (Table 4).

**Table 4.** Number needed to treat and absolute risk reduction with the experimental treatments

| **Treatment** | **NNT** | **ARR, %** | **95% CI of ARR, %** |
| --- | --- | --- | --- |
| TFw | **8** | **12.8** | **-8.7 to 34.4\*** |
| TFd | **4** | **30.8** | **9.7 to 51.9** |

Absolute risk reduction of treatment failure versus the control (terbinafine daily, T).
TFw, terbinafine daily and fluconazole weekly; TFd, terbinafine daily and fluconazole daily; NNT, number needed to treat; ARR, absolute risk reduction; CI, confidence interval.
\*As the 95% CI of ARR extends from a negative number to a positive number, TFw may or may not be superior to control (T).

### Adverse events

One patient in the TFw group was detected to have thrombocytopenia (platelet count <100,000/µl) at 2 weeks and 2 patients in the TFd group were detected to have raised serum glutamic pyruvic transaminase enzyme levels (>100 u/l) at 4 weeks. All 3 patients reported no symptoms due to these derangements. Treatment was stopped in these 3 patients. Test results became normal in subsequent investigations performed after 2 weeks. No other adverse events were detected on investigations and none were reported by the patients.

## Discussion

Present study was conducted in view of unprecedented changes in epidemiology, clinical features and treatment responsiveness of tinea infections in recent past in India.^1-4^ Effectiveness of oral terbinafine,^5, 6^ fluconazole, itraconazole and griseofulvin has decreased considerably, even with 8 weeks of treatment, in recent studies conducted in India.^6^ Furthermore, problems of chronicity as well as frequent relapses occurring within weeks of apparent cure have been noted.^10^ It is obviously important that new treatment methods are tested for their effectiveness in tinea infections.

We tested the hypothesis that using a multidrug therapy regimen may produce better treatment outcome in tinea. Terbinafine and fluconazole were selected as the two antifungal agents for multidrug therapy because they have different mechanisms of action. Effectiveness of two experimental regimens, oral terbinafine daily plus oral fluconazole weekly (TFw) and oral terbinafine daily plus oral fluconazole daily (TFd), was compared with oral terbinafine alone given daily (T, active control) within a randomized pragmatic open design.

Cure rates at 4 weeks with the three treatments were not significantly different. Comparison of cure rates at 8 weeks showed that TFd was more effective than T, while TFw was similar in efficacy to T. Although comparison of cure rates at 8 weeks with TFd (64.1%) and TFw (46.2%) did not reach significance, effect size as calculated by number needed to treat (NNT) (versus terbinafine) showed that TFd was more effective than TFw.

NNT for the experimental group TFw was 8 and for TFd it was 4; meaning thereby that, compared to terbinafine alone, 8 patients need to be treated with TFw to achieve one additional cure, while only 4 patients need to be treated with TFd for the same effect. Thus, TFd was the most effective treatment out of the treatments tested.

Compliance and relapse rates were similar in the three groups. One patient in the TFw group had thrombocytopenia and 2 patients in the TFd group had raised serum glutamic pyruvic transaminase enzyme levels. These changes were reversible, however, it is important to closely monitor liver enzymes with such regimen.

Recent data from randomized pragmatic trials, comparing different drugs and regimens, have shown that itraconazole (5 mg/kg/day) given for 8 weeks had cure rate of 66% in chronic and chronic-relapsing tinea,^6^ and cure rate of 78.3% when all patients with tinea were treated irrespective of the duration of illness.^11^ However, it may be noted that there are limitations with itraconazole, especially with regard to drug interactions and cardiac adverse effects. In such patients, when itraconazole cannot be given, a regimen consisting of terbinafine plus daily fluconazole may be an effective option.

A limitation of the present study is that it was not blinded.

Data presented herein show that it may be worthwhile to consider treatment with oral terbinafine plus daily fluconazole as a treatment option for tinea corporis, tinea cruris and tinea faciei in selected patients in the current epidemic of altered dermatophytosis in India.

## Data Availability

None

